# Development of a self-monitoring system for lung transplant patients using information and communication technology: a pilot study

**DOI:** 10.1101/2023.02.26.23285695

**Authors:** Yoshikazu Shinohara, Kazumichi Yamamoto, Muhammad Wannous, Masahiro Yanagiya, Masaaki Nagano, Kentaro Kitano, Masaaki Sato, Chihiro Konoeda, Jun Nakajima

## Abstract

**Background:** Lung transplantation is the final option for end-stage respiratory diseases. Postoperative monitoring of patients’ physical condition and performance of appropriate interventions for any abnormalities are important to improve the long-term success of lung transplantation. In Japan, patients’ handwritten self-management charts are widely used to record data for home spirometry, vital signs, and medication dosages. However, handwritten data are not suitable for assessment. We developed an internet-based real-time monitoring system (LT-FollowUp) that can easily assess patient data and detect any abnormalities that arise without delay. The aim of this pilot study was to examine the acceptability of LT-FollowUp to patients.

**Methods:** This was a prospective single-arm pilot cohort study. Lung transplant patients were recruited consecutively at regular outpatient visits from October 2020. Patients were instructed to enter their self-measurements (pulmonary functions and vital signs) and immunosuppressant dosages twice a day.

Acceptability was assessed by the data filling rate. The changes in filling rate over time were evaluated using a linear mixed-effects model for repeated measurements.

**Results:** A total of 19 patients were included in the study. There were no significant differences in the filling rates between the handwritten charts and LT-FollowUp.

**Conclusion:** LT-FollowUp is an acceptable system for patients. Further development of the LT-FollowUp system may lead to better long-term clinical outcomes of lung transplantation.

## Introduction

Lung transplantation is the last option for patients with end-stage respiratory failure [1]. Since the enactment of the Organ Transplantation Act in 1997, the number of lung transplantations from neurologically dead donors in Japan has been increasing progressively, including 92 lung transplantations in 2019 [2]. The 5-year survival rates for lung transplantation (58.7% worldwide; 71.2% in Japan) are lower than those for other solid organ transplantations (approximately 90% for heart transplantation and 80% for liver transplantation in Japan) [2, 3, 4]. Chronic rejection and chronic lung allograft dysfunction (CLAD) are the major factors for the poor long-term prognosis of lung transplant patients [5-7].

Postoperative monitoring of patients’ physical condition, medication adherence, and other lung-function-related activities of daily life at the outpatient clinic is important to improve the long-term prognosis after lung transplantation [8]. In particular, home spirometry, a simple respiratory function test that can be performed daily by patients at home, is recommended because it can indicate the presence of CLAD, defined as a decrease in forced expiratory volume in 1 s (FEV1) [9]. Despite the importance of postoperative monitoring, the limited availability of medical personnel and financial resources means that daily home monitoring is largely performed by the patients themselves, with the monitoring data only being checked by the attending physicians at outpatient follow-up visits. Therefore, it is difficult to offer patients adequate care with the optimal timing. In Japan, patients’ handwritten self-management charts are widely used to record the data for home spirometry, other vital signs, and immunosuppressant dosages. However, handwritten data are not suitable for assessment. Furthermore, the handwritten charts are only submitted periodically at regular clinic visits, making it difficult to achieve real-time monitoring and respond rapidly to abnormalities that arise. We developed an internet-based real-time monitoring system that can monitor patients’ data (e.g. home spirometry and vital signs), meaning that physicians can notice any abnormalities even when the patients are at home. Using this system, medical staff and patients can simultaneously notice changes in the patients’ condition using a table format and a graphic visualization. To our knowledge, there is only one comparable digital system for postoperative monitoring of lung transplant patients [10], and our system is the first in Japan. Because this is the first internet-based monitoring system for lung transplant patients, it is important to assess the acceptability of the system to patients and determine the quality of the system.

The aim of this study was to examine the acceptability of our newly developed self-monitoring system (LT-FollowUp) to patients. Acceptability was assessed by the rates of data filling by the patients.

## Methods

### Study design and patients

This was a prospective single-arm pilot cohort study. Patients aged >20 years who had undergone lung transplantation in Japan and attended our outpatient clinic were included. Patients were excluded if they had difficulty in operating electronic devices, had no internet connection, or were unable to provide written consent.

The system was developed in close coordination with the thoracic surgeons who performed the lung transplantations and saw the patients at the outpatient clinic (YS, MY, MN, CK, KK, MS, and JN), a medical doctor who specialized in clinical epidemiology and data science/system development (KY), and a computer scientist who had developed several systems in medical disciplines (WM) [11]. Procedures Patients were recruited at the outpatient clinic because the study started in October 2020. A total of 20 participants were included in the study after providing written consent. The participants received instructions on how to use the LT-FollowUp system. Briefly, they used their own electronic device, such as personal computer, smartphone, or tablet, to connect to a pre-designated URL and log into the system by inputting a provided ID and password. The patients entered their self-measurements (weight, body temperature, heart rate, blood pressure, forced expiratory volume in 6 s [FEV6], FEV1, and arterial oxygen saturation of pulse oximetry [SpO2]) and dosages of immunosuppressants including calcineurin inhibitors (tacrolimus or cyclosporine), antimetabolites (mycophenolate mofetil or azathioprine), and corticosteroids twice a day in the morning and evening (Fig. 1b). FEV6 and FEV1 were measured using the home spirometry system provided to all lung transplant patients. The information entered into LT-FollowUp by the patients was automatically transmitted to our server and the data were simultaneously accessible via the internet to both patients and medical personnel (Fig. 1c). Changes in FEV1, weight, and SpO2 were graphically visualized for easier assessment of trends over time (Fig. 1d). At approximately 1 month after beginning their use of LT-FollowUp, patients were asked about the usability of the system at the outpatient clinic.

**Fig. 1.**
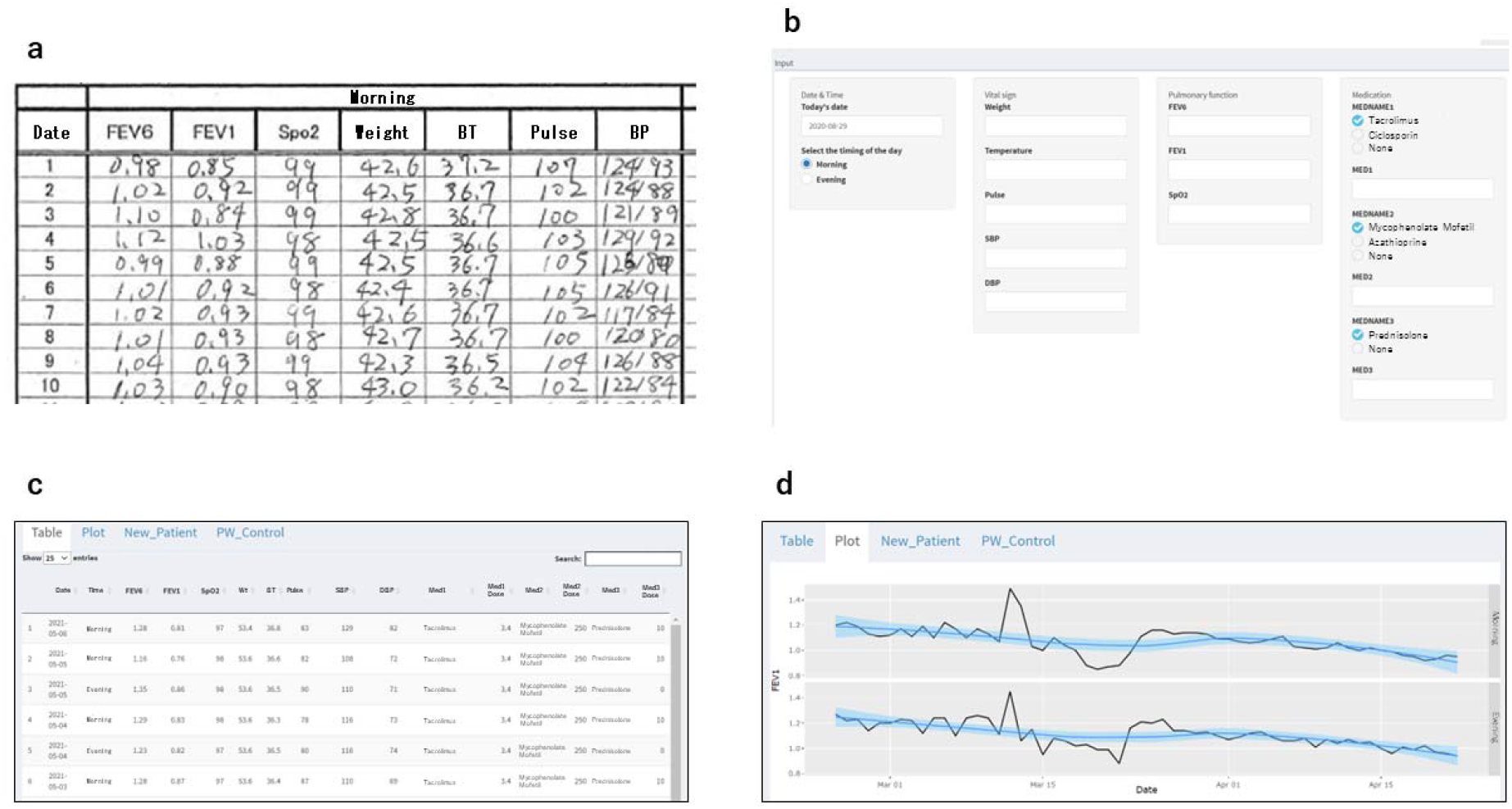
The handwritten self-management chart and screens of LT-FollowUp. (**a)** The handwritten self-management chart used in our hospital. Lung transplant patients log into the system by entering their ID and password. (**b**) The data input screen. Lung transplant patients enter their self-measurements and immunosuppressants. (**c**) The table screen. The input values are listed in the table. (**d**) The graph screen. The input values are visualized by a graph

### Acceptability assessment

The acceptability of LT-FollowUp to the lung transplant patients was assessed by the data filling rate (%), calculated as the number of times that self-monitoring data were actually entered / the total number of opportunities to enter self-monitoring data (twice per day × participation days) × 100. The filling rates were calculated from patients’ handwritten self-management charts (Fig. 1a) for 1 month before the introduction of LT-FollowUp and monthly for 3 months after the introduction of LT-FollowUp (1–30 days, 31–60 days, and 61–90 days).

### Statistical analysis

All statistical analyses were performed using R software v4.0.5 (The R Foundation for Statistical Computing, Vienna, Austria). The changes in filling rate over time were evaluated using a linear mixed-effects model for repeated measures. The model included time (before, 1–30 days, 31–60 days, and 61–90 days) as a fixed effect. The model also included a random intercept for individual patients and a random slope for time to account for within-patient correlations and patient-specific slopes over time. Ethics Statement This study was approved by the Ethics Committee of The University of Tokyo (2020156NI). Written consent was obtained from all participants.

## Results

A total of 20 patients who had undergone lung transplantation at our hospital were recruited for the study. Subsequently, one patient was excluded because of restricted internet access (she was unable to connect to the URL for LT-FollowUp). The remaining 19 patients were included in the analysis. The patient characteristics are shown in Table 1. The largest age group was 40–49 years (48.6 ± 10.7 years). There were more male patients (n = 12) than female patients (n = 7). Idiopathic interstitial pneumonitis was the most common primary disease (n = 6). Most of the participants had undergone lung transplantation at 1–2 years before the study, mainly because the Tokyo Lung Transplant Program did not start until 2015.

**Table 1.**
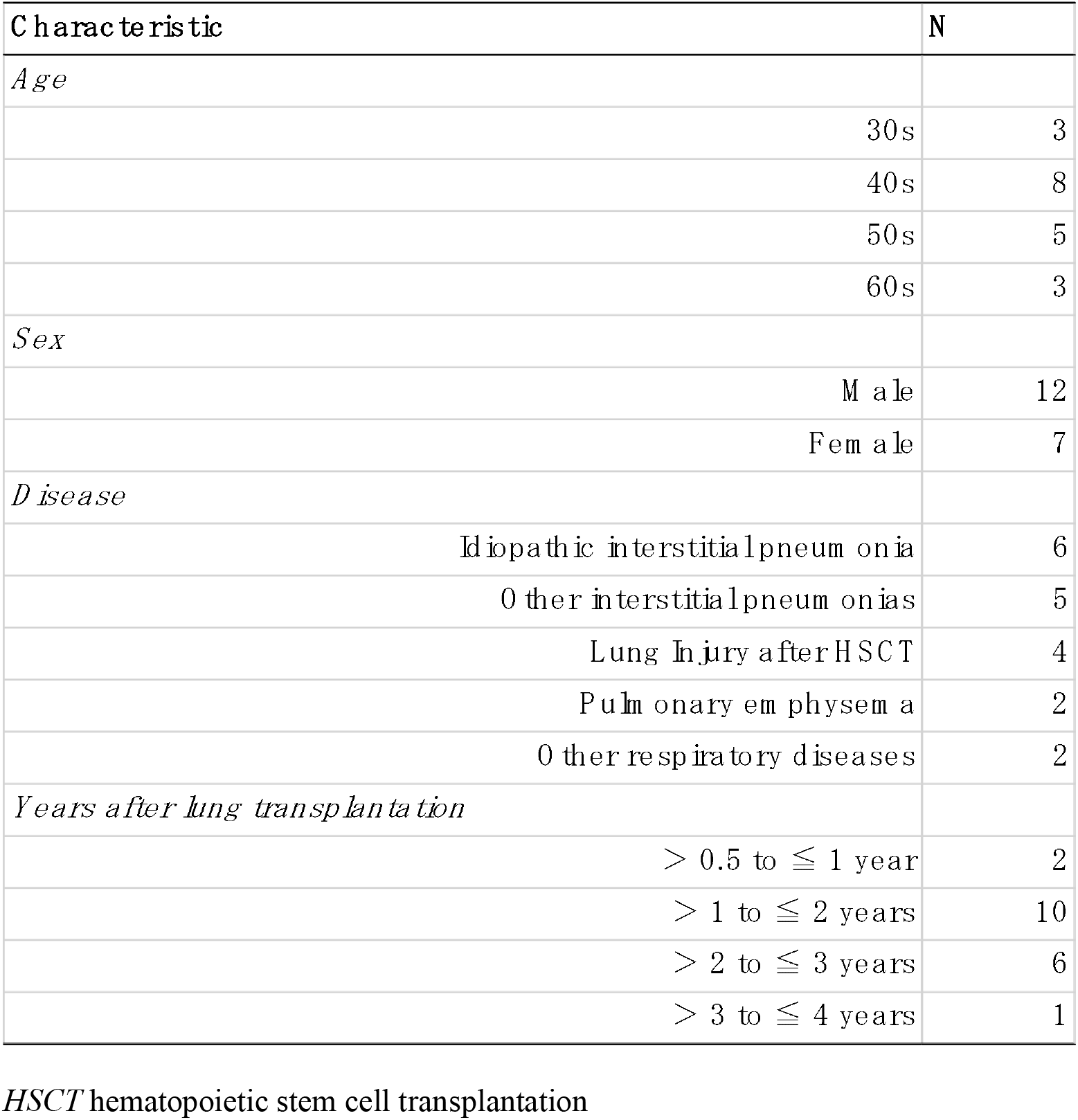
Characteristics of the participating patients (*n* = 19)

The filling rates before and after the introduction of LT-FollowUp are shown in Table 2. Before the introduction of LT-FollowUp, 14 of 19 patients had filling rates >90% on their handwritten charts, and most of the patients performed self-monitoring almost every day. At 1–30, 31–60, and 61–90 days after the introduction of LT-FollowUp, the numbers of patients with filling rates ≥90% were 18, 16, and 16, respectively. There were no significant differences in the filling rates before and after the introduction of LT-FollowUp. Two patients (#8 and #17) showed decreased filling rates over time. Patient #8 performed self-monitoring in the morning only when using the handwritten chart. When she started using LT-FollowUp, she initially entered all required measurements twice a day (morning and evening) as per the protocol. However, for days 61–90, she discontinued the measurements in the evening, meaning that she returned to the same routine used for the handwritten chart. She continued entering all self-monitoring data for LT-FollowUp in the morning. Patient #17 continued to fill in the handwritten chart even after starting LT-FollowUp. He periodically transferred his handwritten records to LT-FollowUp. However, on day 50, he stopped using LT-FollowUp altogether and only continued self-monitoring with the handwritten chart. Patient #14 did not perform regular daily self-monitoring before participation in the study and his filling rate remained very low. We interviewed these three patients (#8, #14, and #17). Patient #14 remained unmotivated about recording his daily measurements in any way throughout the study period, while Patient #17 said that the handwritten records were preferable and he found no further advantage in using the LT-FollowUp system.

**Table 2.** Filling rate before and after use of LT-FollowUp

| Patient # | Age | Sex | Filling rate (%) |  |  |  |
| --- | --- | --- | --- | --- | --- | --- |
|  |  |  | Before the use of LT Follow Up | After the use of LT Follow Up |  |  |
|  |  |  |  | 1-30 days | 31-60 days | 61-90 days |
| 1 | 40s | M | 97.6 | 100.0 | 100.0 | 96.7 |
| 2 | 50s | F | 91.4 | 93.3 | 83.3 | 86.7 |
| 3 | 30s | M | 100.0 | 93.3 | 95.0 | 93.3 |
| 4 | 40s | F | 94.3 | 90.0 | 96.7 | 91.7 |
| 5 | 40s | F | 96.4 | 100.0 | 100.0 | 100.0 |
| 6 | 30s | M | 100.0 | 100.0 | 95.0 | 98.3 |
| 7 | 40s | M | 100.0 | 100.0 | 100.0 | 98.3 |
| 8 | 40s | F | 50.0 | 95.0 | 100.0 | 75.0 |
| 9 | 60s | M | 100.0 | 95.0 | 96.7 | 100.0 |
| 10 | 50s | F | 100.0 | 100.0 | 100.0 | 100.0 |
| 11 | 50s | F | 88.6 | 98.3 | 93.3 | 91.7 |
| 12 | 30s | M | 100.0 | 100.0 | 98.3 | 100.0 |
| 13 | 40s | M | 89.3 | 100.0 | 98.3 | 95.0 |
| 14 | 60s | M | 3.6 | 1.7 | 0.0 | 0.0 |
| 15 | 50s | M | 100.0 | 98.3 | 95.0 | 95.0 |
| 16 | 60s | M | 100.0 | 95.0 | 100.0 | 100.0 |
| 17 | 40s | M | 100.0 | 96.7 | 26.7 | 0.0 |
| 18 | 50s | F | 62.9 | 100.0 | 95.0 | 90.0 |
| 19 | 40s | M | 100.0 | 100.0 | 91.7 | 98.3 |

To estimate the changes in filling rate over time, a linear mixed-effects model for repeated measurements was used. There were no significant changes over time in terms of the filling rates before and after the introduction of LT-FollowUp (beta = −1.50, degrees of freedom (df) = 18, t = −0.721, p = 0.48; Fig. 2).

**Fig. 2.**
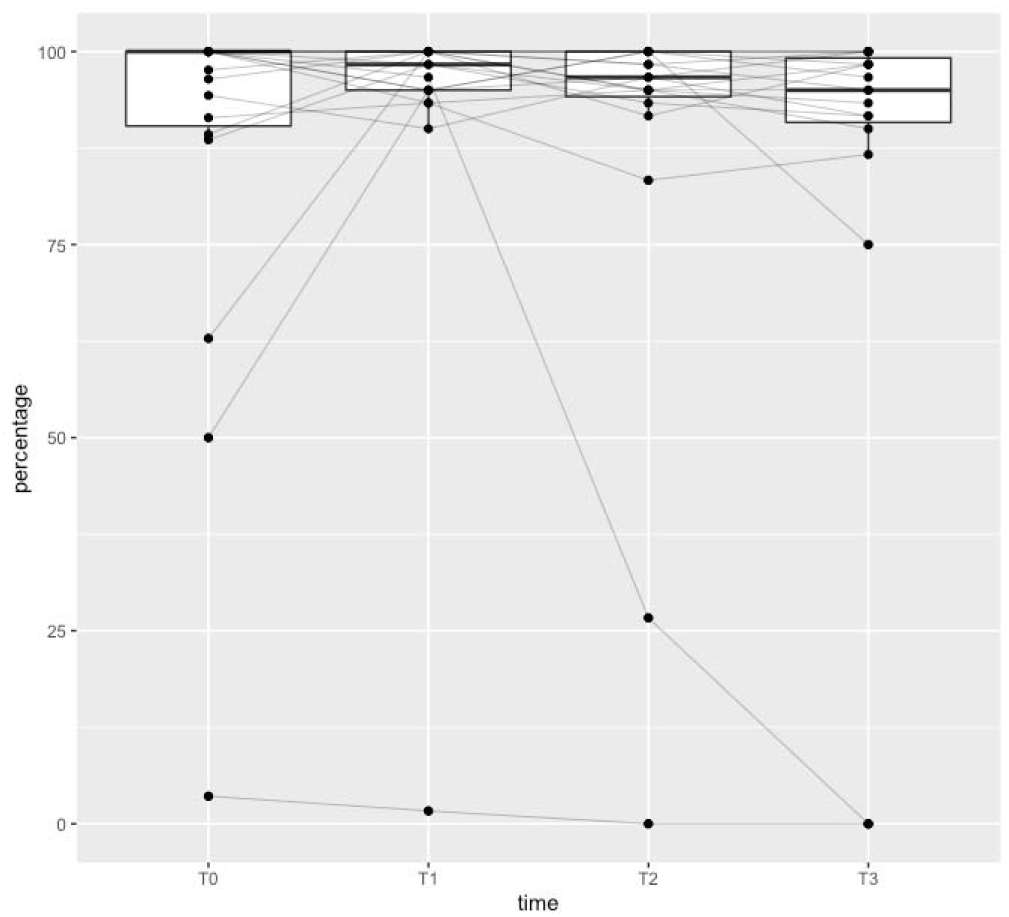
Box plot of the filling rate. The filling rate was calculated monthly after the use of LT-FollowUp. T0: Before use of LT-FollowUp. T1: 1–30 days. T2: 31–60 days. T3: 61–90 days

The LT-FollowUp system did not exhibit any bugs or flaws during the study period, although data entry was disabled for 2 days because of a server problem.

## Discussion

In this study, we developed an internet-based real time-monitoring system to monitor transplant patients’ data (lung function, vital signs, and medication dosages) and detect any abnormalities as early as possible, even when the patients were at home. We evaluated the acceptability of the system to patients by assessing the data filling rates. There were no significant differences in the filling rates before and after the introduction of LT-FollowUp. The overall filling rate for our patients was high before the introduction of LT-FollowUp, and the high filling rate did not decrease after the introduction of LT-FollowUp. Most patients continued to use LT-FollowUp without returning to their handwritten chart, suggesting that the acceptability of the system to patients was satisfactory. Three patients (#8, #14, and #17) showed decreases in their filling rates, suggesting that further modifications and improvements would be indispensable to keep patients motivated.

There are few previous studies related to monitoring applications for lung transplant patients. Dabbs et al. [12] developed pocket PATH, an application for lung transplant patients that runs on Windows mobile devices. For use of pocket PATH, patients needed to borrow a Windows mobile smartphone with pocket PATH pre-installed. In contrast, our LT-FollowUp system can be used on the patients’ own electronic devices by connecting to a pre-designated URL, and thus patients can use their preferred device. This is an advantage of our web-based LT-FollowUp system, because most patients already have a personal electronic device.

In a study using pocket PATH, Jiang et al. [13] reported a decrease in the frequency of pocket PATH use over time: 48% of patients used the application for >75% of days during the first 2 months, 28% of patients used the application for >2 but ≤6 months, and 19% used the application for >6 but ≤12 months. It is possible that the filling rate for the LT-FollowUp system will also decrease over time if we collect data for >3 months.

Some other limitations of the study also require consideration. First, because this was a pilot study, the number of participants was small. Future longer-term studies with more patients are required to assess the long-term outcomes (e.g., filling rates) of LT-FollowUp. Second, we did not evaluate the effect of the real-time monitoring, which is a potential advantage of LT-FollowUp. For example, real-time monitoring for a decrease in FEV1, as an indicator for the presence of CLAD [9], can be conducted by physicians at any time using the LT-FollowUp system. Thus, physicians may be able to note the possible presence of CLAD through real-time LT-FollowUp monitoring before any clinical symptoms emerge. Early detection and immediate initiation of treatment interventions may lead to improvements in the long-term outcomes of lung transplant patients. Development of additional logistics and an alert system is required to realize these benefits of real-time monitoring. This pilot study can act as the basis for future studies designed to improve long-term patient care in the field of lung transplantation by utilizing information and communication technology.

## Conclusion

We have developed an internet-based real-time monitoring system, LT-FollowUp, for lung transplant patients. Patients and physicians can use this system anywhere at any time through their internet devices. In this pilot study, the acceptability of LT-FollowUp to patients was found to be satisfactory. Further development of LT-FollowUp may lead to improvements in the prognosis of lung transplant patients.

## Data Availability

All data produced in the present study are available upon reasonable request to the authors.

## Acknowledgments

The authors thank Alison Sherwin, PhD, from Edanz (https://jp.edanz.com/ac) for editing a draft of this manuscript.

